# *APOE*-ε4 is not associated with pure-tone hearing thresholds, visual acuity or cognition, cross-sectionally or over 3 years of follow up in the Canadian Longitudinal Study on Aging

**DOI:** 10.1101/2023.06.10.23291229

**Authors:** Paul Mick, Rasel Kabir, Malshi Karunatilake, Natalie Phillips, M. Kathleen Pichora-Fuller, Terry-Lyn Young, Yuri Sosero, Ziv Gan-or, Walter Wittich

## Abstract

**Background:** Hearing loss and vision loss are independently and jointly associated with faster rates of cognitive decline. Identifying mechanisms underlying sensory-cognitive associations is a research priority and is needed to inform public health efforts to reduce cognitive decline. Sensory impairment is highly prevalent and treatable, and if a cause-and-effect relationship exists with cognitive decline, treating sensory impairments could reduce rates of cognitive decline with age. On the other hand, if sensory-cognitive associations are the result of a common cause (e.g., a genetic predisposition for both sensory and cognitive impairment), then interventions aimed at reducing sensory loss would not be expected to have beneficial effects on cognition.

The apolipoprotein E (*APOE*)-ε4 allele variant is associated with age-related neurological diseases (e.g., Alzheimer’s disease) and non-neurological diseases (e.g., atherosclerosis).*APOE*-ε4 could be a common factor underlying associations between sensorineural losses and cognitive decline, but links between *APOE*-ε4 and both hearing and vision in the general population remain under-studied. Furthermore, the association between *APOE*-ε4 and cognition in healthy individuals is not as clear as the link between *APOE*-ε4 and Alzheimer’s disease.

Therefore, we aimed to determine if *APOE*-ε4 allele count (the explanatory variable) was associated with differences in baseline and 3-year change in executive function, memory, pure-tone hearing thresholds, and visual acuity (the outcome variables).

**Methods:** A secondary analysis of data collected in the Canadian Longitudinal Study on Aging (CLSA) was performed using data from two time points 3 years apart. Participants, aged 45-85 years, were recruited from 11 cities across Canada. Composite scores for executive function and memory were developed from five tests of cognition. Bilateral air-conduction pure-tone threshold averages and pinhole-corrected visual acuity in the better-seeing eye were used to measure hearing and vision, respectively. Linear mixed regression models assessed associations between *APOE*-ε4 allele count (as a categorical variable with 0 as the reference) and a.) baseline differences and b.) 3-year declines in each of the four outcome variables. Multivariable models adjusted for age, education, sex, race, heart disease, stroke, hypertension and diabetes. Interactions between *APOE*-ε4 and age group (45-54, 55-64, 65-74, and 75-85 years) and *APOE*-ε4 sex were tested.

**Results:** There were 27,765 participants in the CLSA comprehensive cohort but only 11,296 had complete data and were included. Individuals with complete data were more likely to be younger and healthier than those with partially missing data. In main effects models,*APOE*-ε4 was not associated with any of the sensory or cognitive outcome measures, either in terms of differences in baseline values or change over time. Regression models including the*APOE*-ε4*age interaction term (but not the *APOE*-ε4*sex interaction term) better fit the data than the corresponding main effects models. In age-stratified analyses most associations between*APOE*-ε4 and the outcome variables were still not significant. The exceptions were as follows: Twoε4 alleles predicted *better* baseline executive function in the 55-64 year old age group, and *better* baseline pure-tone average in the 45-54 year old age group. In the 65-74 year-old age group, one ε4 allele predicted worsening in visual acuity over time, whereas two ε4 alleles predicted improvements.

**Discussion:** *APOE-*ε4 allele count was not associated with poorer executive function, memory, pure-tone hearing thresholds or visual acuity, at baseline or over 3 years of follow-up, among a population-based sample of healthy 45-85 year old Canadians. Thus, the study does not support the hypothesis that *APOE-*ε4 is a common cause underlying associations between hearing or vision loss (respectively) and declines in each of executive function and memory.

## 1. Introduction

Hearing loss and vision loss are independently and jointly associated with faster rates of cognitive decline (Dearborn et. al., 2018; Ehrlich et al., 2022; Elyashiv et al., 2014; Hämäläinen et al., 2019; Lin et al., 2011; Swenor et al., 2019). Identifying mechanisms underlying sensory-cognitive associations is a research priority and is needed to inform public health efforts to reduce cognitive decline (Nagarajan et al., 2022; Powell, et al., 2022). Sensory impairment is highly prevalent and treatable, and if a cause-and-effect relationship exists with cognitive decline, treating sensory impairments could reduce rates of cognitive decline with age. On the other hand, if sensory-cognitive associations are the result of a common cause (e.g., a genetic predisposition for both sensory and cognitive impairment), then interventions aimed at reducing sensory loss would not be expected to have positive effects on cognition.

The goal of the current study is to determine if the apolipoprotein E (*APOE*) ε4 allele variant is associated with faster declines in visual acuity, hearing, and cognition, to address whether *APOE*-ε4 could be a common cause of sensory-cognitive associations. APOE (note that gene symbols are italicized while protein designations are not)(National Center for Biotechnology Information (US), 2004) is required for a large number of physiological processes that rely on lipid transport (Liu et al., 2013; Mahley and Rall, 2000). The APOE2, APOE3, and APOE4 protein isoforms arise (respectively) from the ε2, ε3, and ε4 allelic variants of the apolipoprotein E (*APOE*) gene The *APOE*-ε4 allele variant is associated with age-related neurological diseases (e.g., Alzheimer’s disease (Liu et al 2013) and dementia with Lewy bodies (Chia et al., 2021)) and non-neurological diseases (e.g., atherosclerosis).

The retina and brain are tissues with similar properties; both are part of the central nervous system, are derived from the same embryonic tissue (Chang et al., 2014; London et al., 2012), and produce APOE (Mahley and Rall, 2000).Associations between vision and cognition have most often been analyzed in healthy samples using visual acuity measures (Nagarajan et al. 2022). Few studies have examined links between APOE and visual acuity in the general population, although some have addressed associations between APOE and specific eye diseases. The APOE4 isoform is associated with cataracts (Utheim et al., 2008; Wu et al., 2015; Zetterberg 2016), open angle glaucoma (Inoue et al., 2013; Paik et al., 2020; Tamura et al., 2006; Wang et al., 2014) and retinal abnormalities such as hard exudates (Santos et al., 2018), venous nicking (Sun et al., 2007) and retinopathy (Liew, 2007). In contrast, the APOE4 protein isoform may be protective against the development of age-related macular degeneration (AMD) (Ishida et al., 2004; Klaver et al., 1998; Levy et al., 2015).

The relationship between *APOE* and hearing loss is not well studied. *APOE*-knockout mice were shown to develop cochlear damage and hearing loss in comparison to wild-type controls (Guo, et al., 2005). In 2012, a population-based study on 435 Dutch residents (85 years and older) found APOE4 to be independently associated with a 2.0 fold increase risk of hearing impairment (Kurniawan et al., 2012), but these results were not replicated in the USA-based Heath, Aging and Body Composition Study (Mener et al., 2016). APOE4 carrier status was not associated with audiometric hearing in a sample of 322 older patients (median age: 71 years) recruited from a hospital in Japan (Morita et al., 2019). The current study is by far the largest to examine associations between hearing loss and APOE

Findings of associations between APOE and cognitive decline in the general population are inconsistent (for a review, see O’Donoghue et al., 2018). Hypothetically, APOE4 might lead to cognitive deficits among individuals who have preclinical, prodromal Alzheimer disease pathology (the “prodromal hypothesis”); alternatively, different isoforms may have differential direct influences on cognition (e.g., via effects on synaptic plasticity and repair), independent of future AD diagnoses (the “phenotype hypothesis”) (Greenwood et al., 2005; Smith et al., 1998). It is possible that differences in study results arise from different ages of participants across different studies.

Antagonistic pleiotropy describes the situation when the influence of a gene changes across the lifespan, being evolutionarily advantageous in early life, but exerting negative effects later in life. The influence of *APOE* on cognition may be more observable in later life (Small et al., 2004), possibly due to gene-related neuronal and neurochemical losses (Lindenberger et al., 2008; McClearn et al., 1997; McGue and Christensen, 2002). Among middle aged individuals, studies have demonstrated that *APOE*-ε4 may be associated with reduced or null cognitive deficits (see Salvato, 2015 for review) or even better cognition (Gharbi-Meliani et al., 2021; Jochemsen et al., 2012).

### 1.1 Objectives

In this study, to address if *APOE*-ε4 could confound sensory-cognitive associations observed in the literature, we aimed to determine if *APOE-*ε4 allele count was associated with cross-sectional and longitudinal differences in a.) b.) memory; c.) pinhole- corrected visual acuity; and d.) pure-tone hearing sensitivity among a large and well-characterized population-based sample of adults who were aged 45-86 years at baseline and who were participating in the Canadian Longitudinal Study on Aging (CLSA)(Raina et al., 2009). To test for evidence of antagonistic pleiotropy, the secondary goal was to to determine if associations were modified by age category (45-54 years, 55-64 years, 65-74 years, and 75-86 years). We also assessed effect modification according to biological sex.

## 2. Materials and Methods

Ethics approval for the secondary analysis of CLSA data was obtained from the University of Saskatchewan Biomedical Research Ethics Board (application ID: 1656) and data access was approved by the CLSA data access committee.

### 2.1. <u>Sample</u>

The CLSA is a population-based closed cohort study with approximately 50,000 participants aged 45-85 years of age recruited from across Canada between 2012-2015. The CLSA is comprised of two cohorts, the tracking and the comprehensive cohorts. Members of the tracking cohort (n∼20,000) respond to telephone-administered questionnaires only, and were excluded from the current analysis. Members of the comprehensive cohort (n∼30,000) provide health information via questionnaire responses, physical examinations (including pure-tone audiometry and visual acuity testing), and analysis of blood samples (including genetic analysis). Thus, we restricted our sample to members of the comprehensive cohort.

Potential CLSA participants are excluded from entry into the study at the time of recruitment if they were judged (by an interviewer) to have cognitive impairment that would interfere with provision of informed consent. Still, 68 of the 30,097 comprehensive cohort members reported a diagnosis of AD at baseline data collection. These individuals were excluded from further analysis. Furthermore, individuals with missing data for any of the variables used in the multivariable models were excluded from the analysis.

### 2.2. Time points

Data were available from the baseline wave of data collection (2012-2015), referred to as “T0” in this manuscript, and from the first wave of follow up (2015-2018), referred to as “T1.” Changes in cognition, hearing, and vision were determined by analyzing changes between T0 and T1. *APOE* genotype and all control variables were measured at T0.

Cross-sectional and longitudinal analyses were performed. In the longitudinal analysis, associations between *APOE*-ε4 allele count (no ε4, ε4 heterozygote (one ε4 allele), ε4 homozygote (two ε4 alleles)) and changes in cognition, pure-tone hearing thresholds and visual acuity over approximately 3 years of follow-up were determined.

### 2.3. Variables of interest

#### 2.3.1. APOE genotype

Each participant’s *APOE* genotype was classified according to number of *APOE* ε4 alleles (0, 1 or 2; 0 being the reference category). Consenting CLSA participants provided venous blood samples at baseline (Canadian Longitudinal Study on Aging, 2015) that were used to determine genotype. DNA extraction and genotyping was performed at the McGill and Genome Québec Innovation Centre in Montréal. Genome-wide genotyping was performed using the Affymetrix UK Biobank Axiom array (Biobank, 2019). The two single nucleotide polymorphisms (SNPs ‒ rs429358 and rs7412) that define the ε2, ε3, and ε4*APOE* genotypes are included in the array. Further details of the CLSA genotyping protocol have been published (Forgetta et al., 2018).

#### 2.3.2. Executive function and memory

Composite cognitive scores for executive function and memory that were previously developed using CLSA data were used in the present analysis (Phillips et al., 2022). Executive function and memory scores were derived from a principal component analysis of the test scores on five cognitive tests: Mental Alternation Test, Animal Fluency test, Controlled Oral Word Association Test, Stroop test, and Rey Auditory Verbal Learning Test with immediate and 5-minute recall. The choice and administration of these tests are described previously by Tuokko et al. (Tuokko et al., 2017). Principal component scores were generated for each time point (T0 and T1).

#### 2.3.3. Bilateral mid-frequency pure-tone threshold average

The primary hearing measure was the bilateral mid-frequency (1000, 2000, 3000, 4000 Hz) pure-tone threshold average (PTA), based on our work showing its superior performance vis-à-vis self-reported hearing loss compared to other summary scores of audiometric thresholds (Mick et al., 2019). Details of audiometry testing in the CLSA have been published (*CLSA Hearing-Audiometer DCS Protocol V3.0 Doc SOP_DCS_0020*, 2014).

#### 2.3.4. Pinhole-corrected visual acuity in the better-seeing eye

The primary measure of vision was the pinhole-corrected visual acuity in the better-seeing eye, based on our previous work showing its superior performance vis-à-vis self-reported vision loss compared to other visual acuity measures used in the CLSA (Mick et al., 2019). Details of visual acuity testing in the CLSA have been published (*CLSA Vision-Visual Acuity Protocol Version 2.2 Document SOP_DCS_0025*, 2017).

### 2.4. Analytic approach

An analysis was performed to describe crude relationships between*APOE-*ε4 allele count and variables of interest. Associations between *APOE*-ε4 allele count and each of the sensory or cognitive variables were then analyzed using repeated measures mixed models. The within-subject effect was defined as time (in days) between data collection site visits (on average about 3 years) for each participant. A multiplicative interaction term between time (as a continuous variable) and ε4 allele count (as a categorical variable) was included in each model (e4 allele count being the between-subject effect of primary interest). Robust estimates of variance were specified in each model (Freedman, 2006). Inverse probability weighting was incorporated into each mixed model using analytic weights calculated by the CLSA to generate parameter and variance estimates more reflective of the general Canadian population in terms of age, sex, province of residence and education level (Canadian Longitudinal Study on Aging, 2020).

#### 2.4.1. Cross-sectional analysis

For each of the dependent variables (executive function, memory, hearing, and vision), predicted mean values (and 95% confidence intervals) at T0 were then calculated for each ε4 allele category (0, 1 or 2 ε4 alleles) using the Stata margins command. Joint tests were performed to assess for statistically significant differences between ε4 allele groups.

#### 2.4.2. Longitudinal analysis

Similarly, for each of the outcome measures, predicted mean annual rate of change (and 95% confidence intervals) were estimated using the Stata margins command. Joint tests were performed to assess for statistically significant differences between ε4 allele groups.

#### 2.4.3. Independent variables in multivariable models

To our knowledge there are no third variables that can be considered common causes of *APOE* genotype and the sensory and cognitive dependent variables. Thus, we did not adjust for confounders in our statistical models (VanderWeele, 2019). We considered forms of selection bias to be potential threats to internal validity because individuals with APOE4-related health problems (that could also affect hearing, vision or cognition) may have been less likely to volunteer to participate in the CLSA (volunteer bias), or be more likely to drop out of the study (attrition bias). Adjusting for such conditions, however, could introduce bias if such conditions actually mediate relationships between*APOE* genotype and the outcomes of interest (Hernán et al., 2004). To address the dilemma, we performed both crude and multivariable analyses. In the multivariable analyses, we adjusted for age (as a linear term), education level, sex, white race/ethnicity, and histories of angina, myocardial infarction, stroke, hypertension, and diabetes. In models examining associations between *APOE*-ε4 allele count and cognition, we also adjusted for PTA and visual acuity; and in models examining associations between *APOE*-ε4 allele count and sensory outcomes, we also adjusted for principal component scores for executive function and memory.

#### 2.4.4. Interaction models for age and sex

After each multivariable main effect model was run, effect modification according to 4-category age category at baseline (45-54, 55-64, 65-74, 75-86 years) was analyzed. For each participant, each of the two (time× ε4 allele count) indicator variables used in the main effects model were further multiplied by each of the three age category indicator variables (age 45-54 was the referent category). The interaction between age category and ε4 allele count on change in the dependent variable over time was considered significant if the Akaike Information Criterion (AIC) value for the more complex model was lower than the AIC value for the nested simpler model (Bozdogam, 1987). Effect modification according to sex was then analyzed in an analogous way.

Sex was determined by the question (asked at baseline), “Are you male or female?” The CLSA included a gender identity questionnaire at T1 that recorded transgender responses. Comparing the responses to the sex and gender variables indicated that >99.5% of participants were cis-gendered. Furthermore, gender (rather than sex)-influenced factors are not known to affect *APOE* expression and thus used sex rather than gender in the analysis of interaction.

## 3. Results

### 3.1. Sample characteristics (*Table 1*)

The sample included 11,296 participants who had complete data for all of the variables used in the multivariable models; 8,335 participants (73.8%) had noε4 alleles, 2,746 (24.3%) had one ε4 allele, and 215 (1.9%) had two ε4 alleles. The demographic and health characteristics of people grouped according to number of ε4 alleles (0, 1 or 2) were similar. There were slight differences in the proportion of people with diabetes and the proportion of people in different income categories across ε4 groups (Table 1).

**Table 1.**
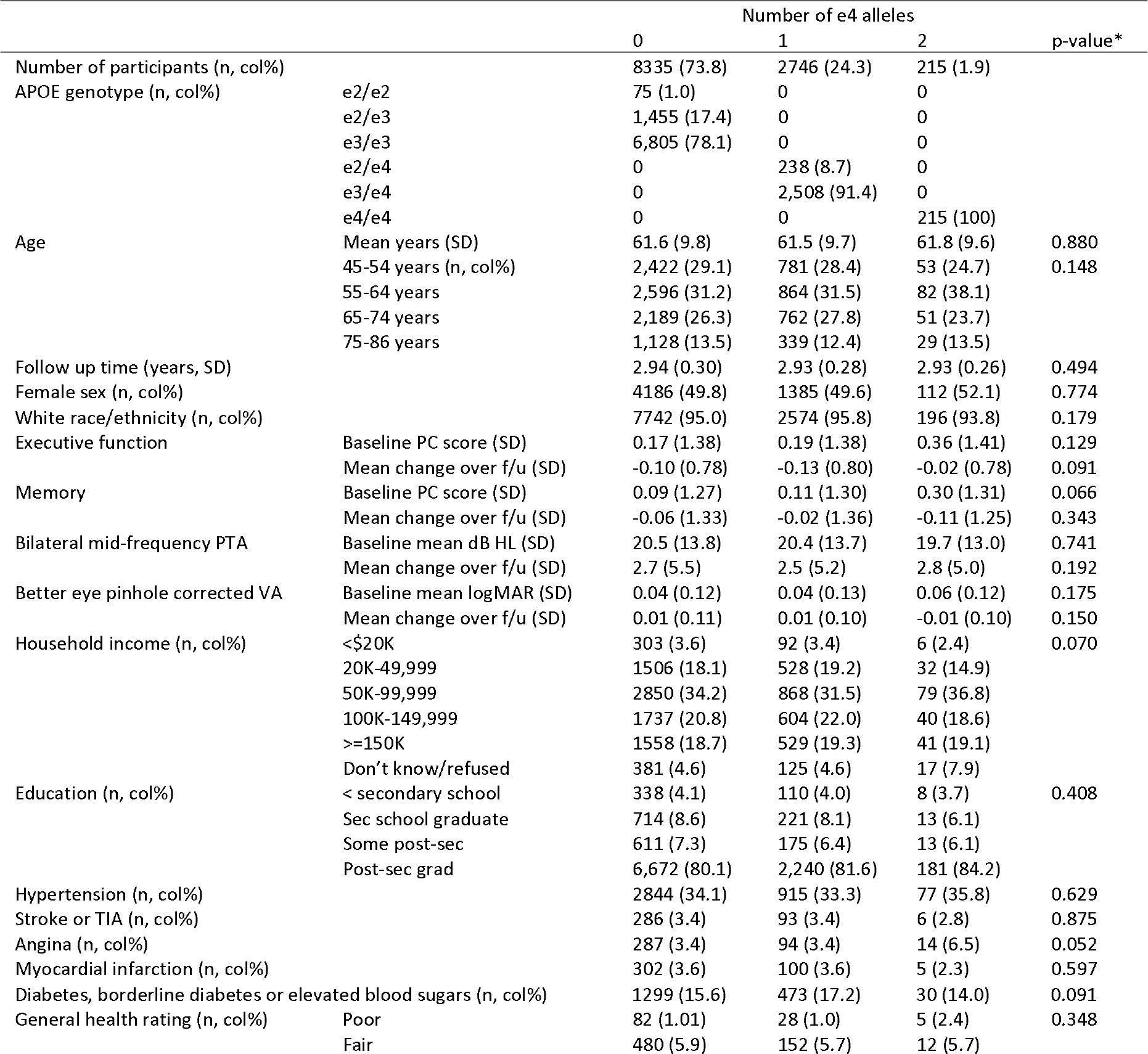
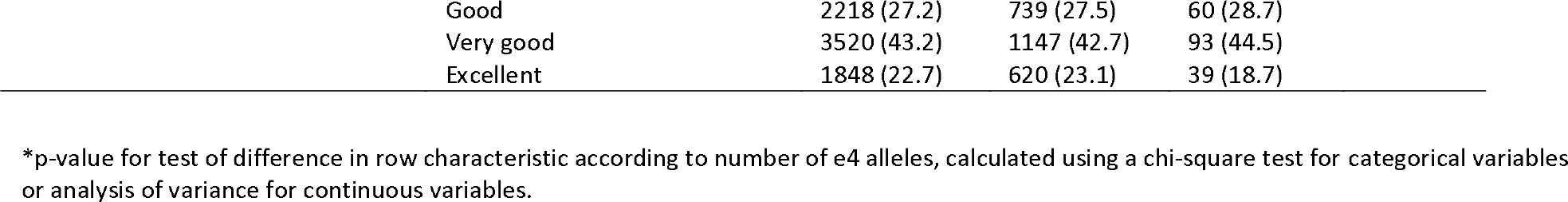
Descriptive statistics of individuals included in the analysis (baseline data)

|  |  | Number of e4 alleles |  |  | p-value* |
| --- | --- | --- | --- | --- | --- |
|  |  | 0 | 1 | 2 |  |
| Number of participants (n, col%) |  | 8335 (73.8) | 2746 (24.3) | 215 (1.9) |  |
| APOE genotype (n, col%) | e2/e2 | 75 (1.0) | 0 | 0 |  |
|  | e2/e3 | 1,455 (17.4) | 0 | 0 |  |
|  | e3/e3 | 6,805 (78.1) | 0 | 0 |  |
|  | e2/e4 | 0 | 238 (8.7) | 0 |  |
|  | e3/e4 | 0 | 2,508 (91.4) | 0 |  |
|  | e4/e4 | 0 | 0 | 215 (100) |  |
| Age | Mean years (SD) | 61.6 (9.8) | 61.5 (9.7) | 61.8 (9.6) | 0.880 |
|  | 45-54 years (n, col%) | 2,422 (29.1) | 781 (28.4) | 53 (24.7) | 0.148 |
|  | 55-64 years | 2,596 (31.2) | 864 (31.5) | 82 (38.1) |  |
|  | 65-74 years | 2,189 (26.3) | 762 (27.8) | 51 (23.7) |  |
|  | 75-86 years | 1,128 (13.5) | 339 (12.4) | 29 (13.5) |  |
| Follow up time (years, SD) |  | 2.94 (0.30) | 2.93 (0.28) | 2.93 (0.26) | 0.494 |
| Female sex (n, col%) |  | 4186 (49.8) | 1385 (49.6) | 112 (52.1) | 0.774 |
| White race/ethnicity (n, col%) |  | 7742 (95.0) | 2574 (95.8) | 196 (93.8) | 0.179 |
| Executive function | Baseline PC score (SD) | 0.17 (1.38) | 0.19 (1.38) | 0.36 (1.41) | 0.129 |
|  | Mean change over f/u (SD) | -0.10 (0.78) | -0.13 (0.80) | -0.02 (0.78) | 0.091 |
| Memory | Baseline PC score (SD) | 0.09 (1.27) | 0.11 (1.30) | 0.30 (1.31) | 0.066 |
|  | Mean change over f/u (SD) | -0.06 (1.33) | -0.02 (1.36) | -0.11 (1.25) | 0.343 |
| Bilateral mid-frequency PTA | Baseline mean dB HL (SD) | 20.5 (13.8) | 20.4 (13.7) | 19.7 (13.0) | 0.741 |
|  | Mean change over f/u (SD) | 2.7 (5.5) | 2.5 (5.2) | 2.8 (5.0) | 0.192 |
| Better eye pinhole corrected VA | Baseline mean logMAR (SD) | 0.04 (0.12) | 0.04 (0.13) | 0.06 (0.12) | 0.175 |
|  | Mean change over f/u (SD) | 0.01 (0.11) | 0.01 (0.10) | -0.01 (0.10) | 0.150 |
| Household income (n, col%) | <\$20K | 303 (3.6) | 92 (3.4) | 6 (2.4) | 0.070 |
|  | 20K-49,999 | 1506 (18.1) | 528 (19.2) | 32 (14.9) |  |
|  | 50K-99,999 | 2850 (34.2) | 868 (31.5) | 79 (36.8) |  |
|  | 100K-149,999 | 1737 (20.8) | 604 (22.0) | 40 (18.6) |  |
|  | >=150K | 1558 (18.7) | 529 (19.3) | 41 (19.1) |  |
|  | Don't know/refused | 381 (4.6) | 125 (4.6) | 17 (7.9) |  |
| Education (n, col%) | < secondary school | 338 (4.1) | 110 (4.0) | 8 (3.7) | 0.408 |
|  | Sec school graduate | 714 (8.6) | 221 (8.1) | 13 (6.1) |  |
|  | Some post-sec | 611 (7.3) | 175 (6.4) | 13 (6.1) |  |
|  | Post-sec grad | 6,672 (80.1) | 2,240 (81.6) | 181 (84.2) |  |
| Hypertension (n, col%) |  | 2844 (34.1) | 915 (33.3) | 77 (35.8) | 0.629 |
| Stroke or TIA (n, col%) |  | 286 (3.4) | 93 (3.4) | 6 (2.8) | 0.875 |
| Angina (n, col%) |  | 287 (3.4) | 94 (3.4) | 14 (6.5) | 0.052 |
| Myocardial infarction (n, col%) |  | 302 (3.6) | 100 (3.6) | 5 (2.3) | 0.597 |
| Diabetes, borderline diabetes or elevated blood sugars (n, col%) |  | 1299 (15.6) | 473 (17.2) | 30 (14.0) | 0.091 |
| General health rating (n, col%) | Poor | 82 (1.01) | 28 (1.0) | 5 (2.4) | 0.348 |
|  | Fair | 480 (5.9) | 152 (5.7) | 12 (5.7) |  |

|  |  |  |  |
| --- | --- | --- | --- |
| Good | 2218 (27.2) | 739 (27.5) | 60 (28.7) |
| Very good | 3520 (43.2) | 1147 (42.7) | 93 (44.5) |
| Excellent | 1848 (22.7) | 620 (23.1) | 39 (18.7) |

\*p-value for test of difference in row characteristic according to number of e4 alleles, calculated using a chi-square test for categorical variables or analysis of variance for continuous variables.

### 3.2. Allele frequency

*APOE* allele frequency was calculated to compare against other samples in the literature. There were 22,592 alleles counted in the sample (11,296 participants x 2*APOE* alleles/participant). There were 1,843 ε2 alleles (8.2%), 17,573 ε3 alleles (77.8%) and 3,176 ε4 alleles (14.1%).

### 3.3. Missing data (*Table 2*)

There were 27,765 participants in the CLSA comprehensive cohort who participated in both waves of data collection. Of these individuals, 9,558 did not have genetic information available (34.4%), 10,578 (38.1%) had missing cognition data, 3,748 (13.5%) had missing audiometric data, and 2,239 (8.1%) had missing visual acuity data. Very little data were missing for other variables, which were mostly questionnaire items. Some participants were missing data for more than one variable. Table 2 compares the characteristics of participants with complete data, who were included in the analysis, with participants who were missing data for at least one of the variables and who were excluded from the analysis. Compared to participants with missing data, participants with complete data were younger (mean 61.6 years versus 63.4 years), slightly less likely to be female (49.8% versus 51.6%), and had better executive function (mean baseline principal component score -0.14 versus 0.18), memory (mean baseline principal component score -0.08 versus 0.10), pure-tone hearing (mean baseline PTA 23.2 versus 20.4 dB HL), and visual acuity (mean baseline score of 0.07 versus 0.04 logMAR). They were slightly more likely to have higher income and education, and were less likely to have histories of hypertension, stroke or transient ischemic attack, myocardial infarction, or diabetes. Their self-ratings of general health were slightly better, as well. There were no significant differences, however, in e4 allele profiles between participants with complete data versus those with missing data.

**Table 2.** Comparison of characteristics between CLSA participants with complete data for the variables used in the analysis (who were included in the analysis) and participants with missing data (who were excluded).

|  |  | Missing data;<br>Excluded<br>from analysis | Complete<br>data;<br>included in<br>analysis | P-value* |
| --- | --- | --- | --- | --- |
| Number of participants (n, col%) |  | 16,469 (59.3) | 11,296 (99.4) |  |
| Number of e4 alleles (n, col%) | 0 | 6,253 (74.8) | 8,335 (73.8) | 0.270 |
|  | 1 | 1,950 (23.3) | 2,746 (24.3) |  |
|  | 2 | 161 (1.9) | 215 (1.9) |  |
| Genotype (n, col%) | e2/e2 | 51 (0.6) | 75 (0.7) | 0.602 |
|  | e2/e3 | 1,062 (12.7) | 1,455 (12.9) |  |
|  | e3/e3 | 5,140 (61.5) | 6,805 (60.2) |  |
|  | e2/e4 | 175 (2.1) | 238 (2.1) |  |
|  | e3/e4 | 1,775 (21.2) | 2,508 (22.2) |  |
|  | e4/e4 | 161 (1.9) | 215 (1.9) |  |
| Age at baseline | Mean years (SD) | 63.9 (10.4) | 61.6 (9.8) | <0.001 |
|  | 45-54 years (n, col%) | 4,339 (23.1) | 3,256 (28.8) | <0.001 |
|  | 55-64 years | 5,297 (28.2) | 3,542 (31.4) |  |
|  | 65-74 years | 5,377 (28.6) | 3,002 (26.6) |  |
|  | 75-86 years | 3,788 (20.2) | 1,496 (13.2) |  |
| Follow up time (years, SD) |  | 2.98 (0.37) | 2.93 (0.30) | <0.001 |
| Female sex (n, col%) |  | 9,698 (51.6) | 5,622 (49.8) | 0.002 |
| White race/ethnicity (n, col%) |  | 17,630 (93.8) | 10,742 (95.1) | <0.001 |
| Executive function | Baseline PC score (SD) | -0.14 (1.50) | 0.18 (1.38) | <0.001 |
|  | Change over f/u | -0.12 (0.82) | -0.11 (0.79) | 0.495 |
| Memory | Baseline PC score (SD) | -0.08 (1.35) | 0.10 (1.28) | <0.001 |
|  | Change over f/u | -0.07 (1.35) | -0.05 (1.33) | 0.349 |
| Mid-frequency PTA | Baseline mean dB HL (SD) | 23.2 (15.3) | 20.4 (13.7) | <0.001 |
|  | Change over f/u | 2.9 (5.7) | 2.6 (5.4) | <0.001 |
| Better eye pinhole corrected VA | Baseline mean logMAR (SD) | 0.07 (0.14) | 0.04 (0.13) | <0.001 |
|  | Change over f/u | 0.007 (0.111) | 0.006 (0.106) | 0.426 |
| Household income (n, col%) | <\$20K | 1,165 (6.2) | 401 (3.6) | <0.001 |
|  | 20K-49,999 | 4,294 (22.8) | 2,066 (18.3) |  |
|  | 50K-99,999 | 6,110 (32.5) | 3,797 (33.6) |  |
|  | 100K-149,999 | 3,143 (16.7) | 2,381 (21.1) |  |
|  | >=150K | 2,671 (14.20) | 2,128 (18.8) |  |
|  | Don't know/refused | 1,418 (7.5) | 523 (4.6) |  |
| Education (n, col%) | < secondary school | 1,187 (6.3) | 456 (4.0) | <0.001 |
|  | Sec school graduate | 1,891 (10.1) | 948 (8.4) |  |
|  | Some post-sec | 1,439 (7.7) | 799 (7.1) |  |
|  | Post-sec grad | 14,234 (75.9) | 9,093 (80.5) |  |
| Hypertension (n, col%) |  | 7,265 (39.0) | 3,836 (34.0) | <0.001 |
| Stroke or TIA (n, col%) |  | 962 (5.2) | 385 (3.4) | <0.001 |
| Angina (n, col%) |  | 929 (5.0) | 395 (3.5) | <0.001 |
| Myocardial infarction (n, col%) |  | 1,054 (5.7) | 407 (3.6) | <0.001 |
| Diabetes, borderline diabetes or elevated blood sugars (n, col%) |  | 3,508 (18.8) | 1,802 (16.0) | <0.001 |
| General health rating (n, col%) | Poor | 3,460 (18.4) | 2,535 (22.5) | <0.001 |
|  | Fair | 7,555 (40.2) | 4,865 (43.1) |  |
|  | Good | 5,770 (30.7) | 3,107 (27.5) |  |
|  | Very good | 1,651 (8.8) | 664 (5.9) |  |
|  | Excellent | 347 (1.9) | 120 (1.1) |  |

\*p-value for test of difference in row characteristic according to missingness (complete vs. missing), calculated using a chi-square test for categorical variables or 2-tailed t-test for continuous variables

### 3.4. Associations between APOE ε4 allele count and outcomes of interest (*Tables 3 and 4*)

In main effects models, *APOE-*ε4 allele count was not significantly associated with any of the sensory or cognitive outcome measures, at baseline or in terms of rate of change.In each case, however, the model with the APOE*age interaction term better fit the data than corresponding main effects model (in contrast, the main effects models better fit the data than the corresponding model with the APOE*sex interaction term). Thus, we used regression models that included an APOE*age interaction term, and present results stratified by age groups (45- 54, 55-64, 65-74, 75-85 years old). There were no meaningful differences in results between crude and multivariable models, and only the results for multivariable models are reported in the text of main manuscript. Results for the crude models can be found in Tables 5 (baseline analyses) and 6 (longitudinal analyses).

**Table 3.**
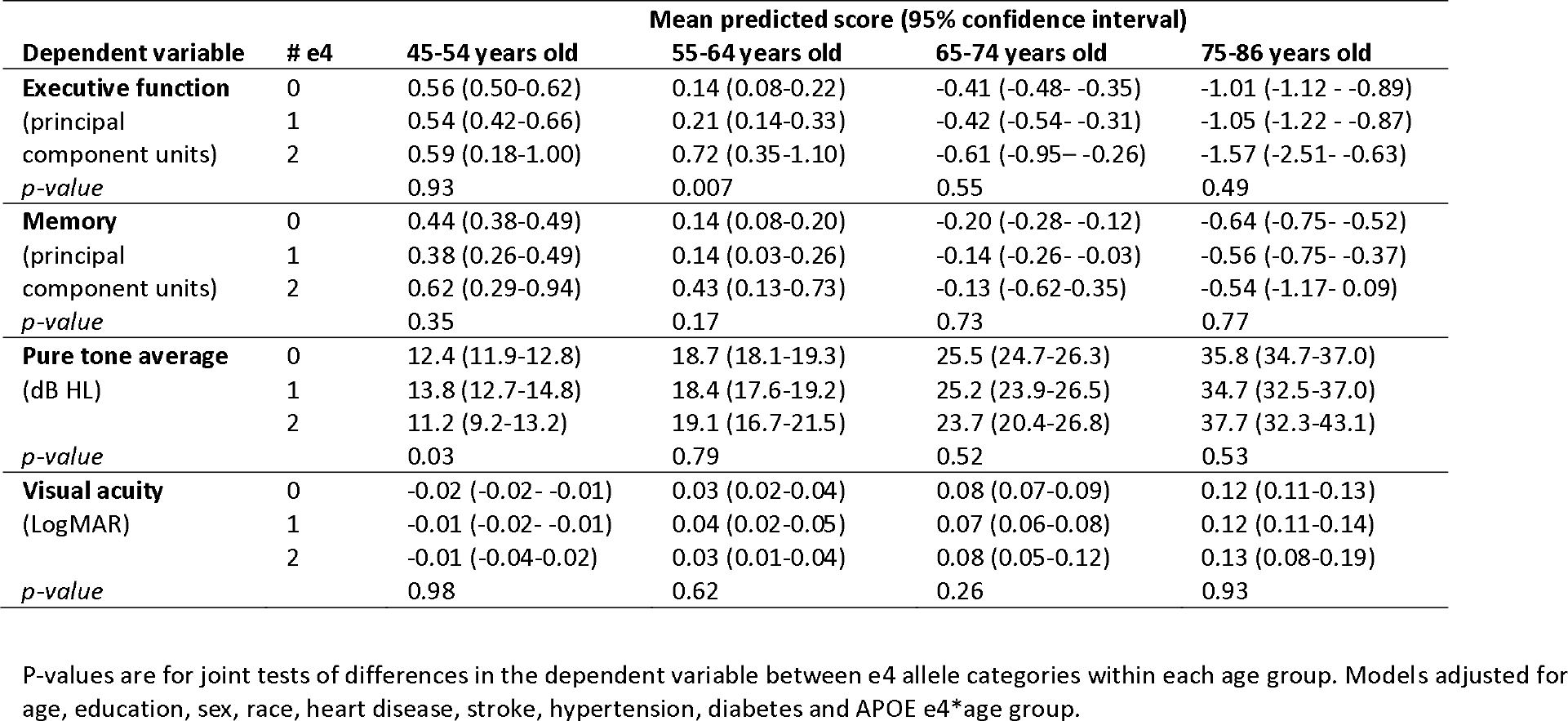
Predicted average cognitive and sensory scores at baseline for participants with 0, 1 and 2 *APOE* e4 alleles, calculated from multivariable linear mixed regression models.

| Dependent variable | # e4 | Mean predicted score (95% confidence interval) |  |  |  |
| --- | --- | --- | --- | --- | --- |
|  |  | 45-54 years old | 55-64 years old | 65-74 years old | 75-86 years old |
| **Executive function**<br>(principal component units) | 0 | 0.56 (0.50-0.62) | 0.14 (0.08-0.22) | -0.41 (-0.48- -0.35) | -1.01 (-1.12 - -0.89) |
|  | 1 | 0.54 (0.42-0.66) | 0.21 (0.14-0.33) | -0.42 (-0.54- -0.31) | -1.05 (-1.22 - -0.87) |
|  | 2 | 0.59 (0.18-1.00) | 0.72 (0.35-1.10) | -0.61 (-0.95- -0.26) | -1.57 (-2.51- -0.63) |
| *p-value* |  | 0.93 | 0.007 | 0.55 | 0.49 |
| **Memory**<br>(principal component units) | 0 | 0.44 (0.38-0.49) | 0.14 (0.08-0.20) | -0.20 (-0.28- -0.12) | -0.64 (-0.75- -0.52) |
|  | 1 | 0.38 (0.26-0.49) | 0.14 (0.03-0.26) | -0.14 (-0.26- -0.03) | -0.56 (-0.75- -0.37) |
|  | 2 | 0.62 (0.29-0.94) | 0.43 (0.13-0.73) | -0.13 (-0.62-0.35) | -0.54 (-1.17- 0.09) |
| *p-value* |  | 0.35 | 0.17 | 0.73 | 0.77 |
| **Pure tone average**<br>(dB HL) | 0 | 12.4 (11.9-12.8) | 18.7 (18.1-19.3) | 25.5 (24.7-26.3) | 35.8 (34.7-37.0) |
|  | 1 | 13.8 (12.7-14.8) | 18.4 (17.6-19.2) | 25.2 (23.9-26.5) | 34.7 (32.5-37.0) |
|  | 2 | 11.2 (9.2-13.2) | 19.1 (16.7-21.5) | 23.7 (20.4-26.8) | 37.7 (32.3-43.1) |
| *p-value* |  | 0.03 | 0.79 | 0.52 | 0.53 |
| **Visual acuity**<br>(LogMAR) | 0 | -0.02 (-0.02- -0.01) | 0.03 (0.02-0.04) | 0.08 (0.07-0.09) | 0.12 (0.11-0.13) |
|  | 1 | -0.01 (-0.02- -0.01) | 0.04 (0.02-0.05) | 0.07 (0.06-0.08) | 0.12 (0.11-0.14) |
|  | 2 | -0.01 (-0.04-0.02) | 0.03 (0.01-0.04) | 0.08 (0.05-0.12) | 0.13 (0.08-0.19) |
| *p-value* |  | 0.98 | 0.62 | 0.26 | 0.93 |

P-values are for joint tests of differences in the dependent variable between e4 allele categories within each age group. Models adjusted for age, education, sex, race, heart disease, stroke, hypertension, diabetes and *APOE* e4\*age group.

**Table 4.** Predicted average change in cognitive and sensory scores from baseline to 3-year follow up for participants with 0, 1 and 2 e4 alleles, calculated from multivariable linear mixed regression models.

| Dependent variable | # e4 | Mean predicted change in score (95% CI) |  |  |  |
| --- | --- | --- | --- | --- | --- |
|  |  | 45-54 years old | 55-64 years old | 65-74 years old | 75-86 years old |
| **Executive function**<br>(principal component units) | 0 | -0.02 (-0.03- -0.006) | -0.02 (-0.04- -0.01) | -0.05 (-0.06- -0.03) | -0.05 (-0.08- -0.03) |
|  | 1 | -0.02 (-0.05 - 0.004) | -0.03 (-0.05 - -0.01) | -0.05 (-0.08- -0.02) | -0.10 (-0.15 - -0.05) |
|  | 2 | 0.01 (-0.08 - 0.10) | -0.02 (-0.07 - 0.04) | 0.02 (-0.08 - 0.11) | 0.05 (-0.14 - 0.24) |
|  | *p-value* | 0.82 | 0.79 | 0.43 | 0.29 |
| **Memory**<br>(principal component units) | 0 | 0.01 (-0.01-0.03) | -0.01 (-0.03-0.02) | -0.04 (-0.06- -0.01) | -0.09 (-0.13- -0.04) |
|  | 1 | 0.03 (-0.01-0.07) | -0.01 (-0.05-0.03) | -0.03 (-0.08-0.02) | -0.03 (-0.11-0.05) |
|  | 2 | -0.002 (-0.09-0.09) | -0.01 (-0.10-0.08) | -0.04 (-0.20-0.12) | -0.19 (-0.39-0.01) |
|  | *p-value* | 0.64 | 0.99 | 0.99 | 0.28 |
| **Pure tone average**<br>(dB HL) | 0 | 0.57 (0.50-0.64) | 0.82 (0.71-0.93) | 1.04 (0.92-1.16) | 1.47 (1.30-1.65) |
|  | 1 | 0.45 (0.27-0.62) | 0.79 (0.66-0.92) | 1.07 (0.92-1.22) | 1.46 (1.25-1.67) |
|  | 2 | 0.39 (-0.17-0.95) | 0.94 (0.53-1.36) | 0.76 (0.37-1.16) | 1.57 (0.19-2.95) |
|  | *p-value* | 0.36 | 0.79 | 0.37 | 0.99 |
| **Visual acuity**<br>(logMAR) | 0 | 0.004 (-0.001-0.008) | 0.002 (0.000-0.004) | 0.001 (-0.001-0.003) | 0.004 (0.001-0.008) |
|  | 1 | 0.005 (-0.002-0.012) | 0.000 (-.003-0.004) | 0.006 (0.001-0.011) | 0.005 (-0.002-0.012) |
|  | 2 | -0.002 (-.026-0.022) | -0.004 (-0.010- .003) | -0.013 (-.024- -.002) | -0.002 (-.026-0.022) |
|  | *p-value* | 0.82 | 0.19 | 0.006 | 0.82 |

P-values are for joint tests of differences in the dependent variable between e4 allele categories within each age group. Models adjusted for age, education, sex, race, heart disease, stroke, hypertension, diabetes, and APOE e4\*age group.

**Table 5.**
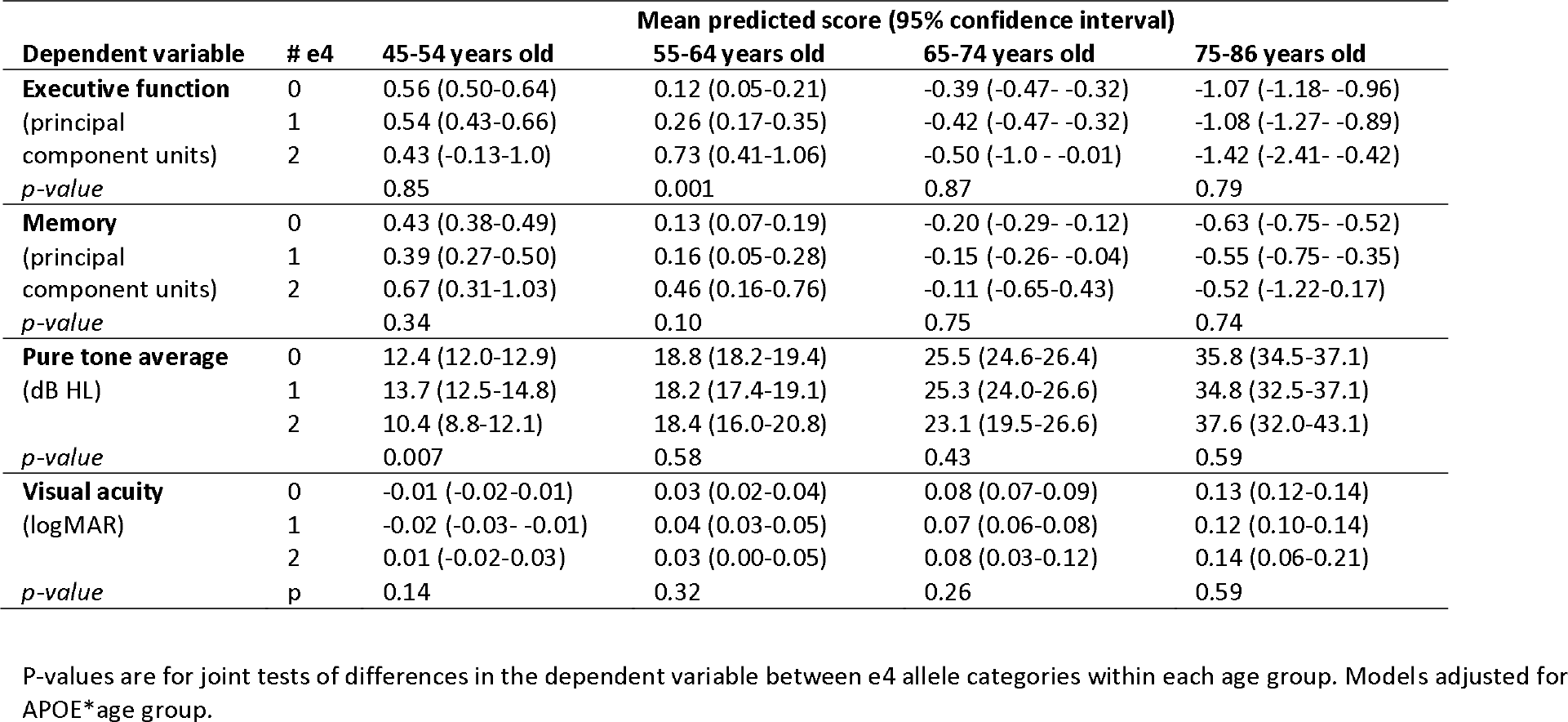
Predicted average cognitive and sensory scores at baseline for participants with 0, 1 and 2 *APOE* e4 alleles, calculated from crude linear mixed regression models.

**Table 6.**
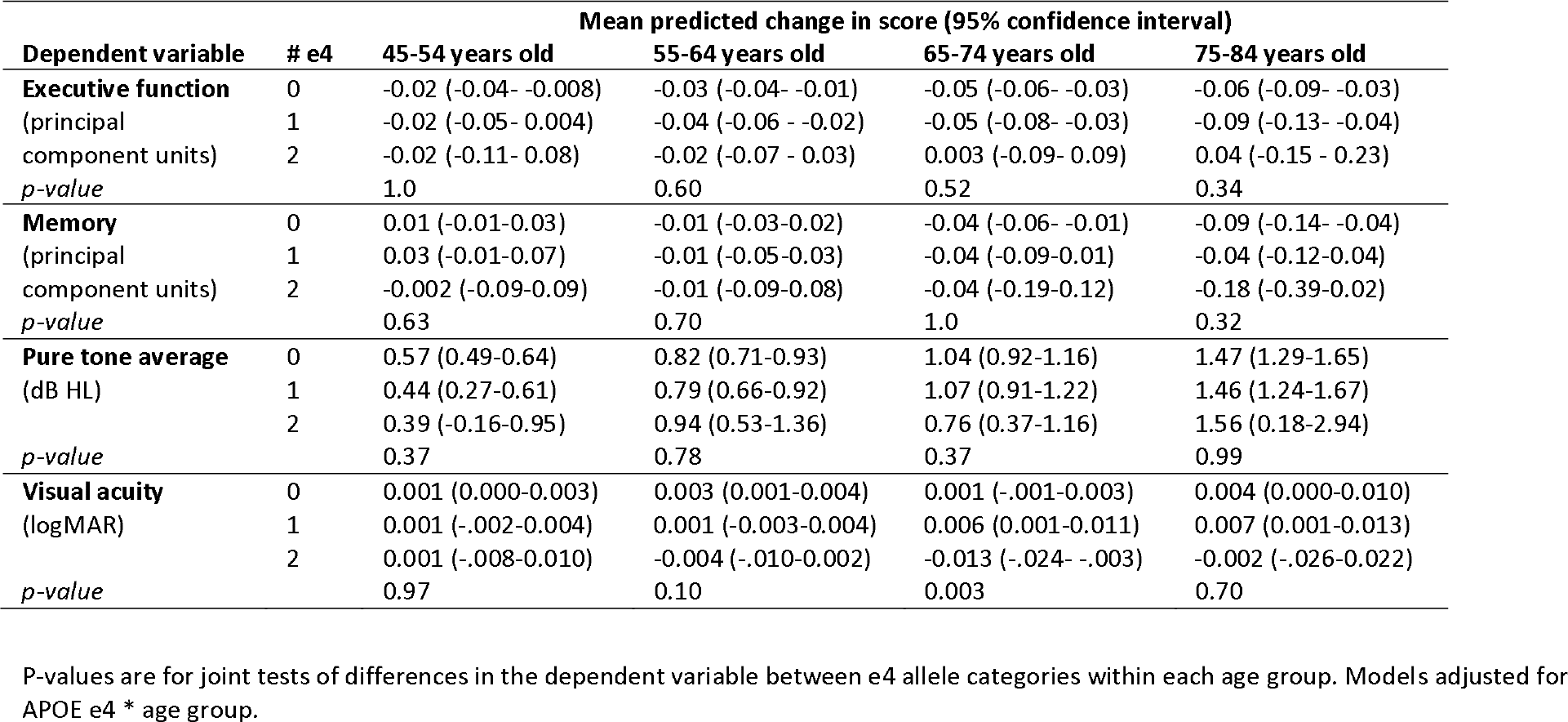
Predicted average change in cognitive and sensory scores from baseline to 3-year follow up for participants with 0, 1 and 2 *APOE* e4 alleles, calculated from crude linear mixed regression models.

#### 3.4.1 Executive function

In the 55-64-year-old age group, individuals with two ε4 alleles had *better* executive function scores at baseline than individuals with no or just oneε4 allele. There were no differences in executive function scores between ε4 groups in other age groups (Table 3). There were no differences in rate of change in executive function over time betweenε4 groups in any of the age groups (table 4).

#### 3.4.2. Memory

There were no differences in memory scores at baseline, or in change in memory scores over the follow up period, between ε4 groups in any of the age groups (Tables 3 and 4).

#### 3.4.3. Hearing

At baseline, among 45-54-year-olds, binaural mid-frequency PTA was slightly *better* for ε4 homozygotes than other ε4 groups (Table 3). No other differences (cross-sectionally or longitudinally) between ε4 groups were observed in any age group (Tables 3 and 4).

#### 3.4.4. Vision

No differences in better-eye pinhole-corrected visual acuity were observed between ε4 groups at baseline for any of the age groups. Among 65-74-year-olds, change in visual acuity over time differed between ε4 groups; it slightly improved for *APOE* ε4 homozygotes, whereas it slightly worsened for ε4 heterozygotes, and stayed the same for individuals with no ε4 alleles (Table 4).

## 4. Discussion

The results do not support the hypothesis that *APOE* ε4 allele count increases the risk of having or developing poorer executive function, memory, pure-tone hearing or visual acuity among a population-based sample of healthy adults aged 45-85 years at baseline. Against expectations, in some age groups, *APOE* ε4 homozygotes had better outcomes than other participants.

Specifically, among 45-54-year-olds, they had better hearing; among 55-64-year-olds they had better executive function, and among 65-74-year-olds their visual acuity slightly improved over time. The age-specific results should be considered exploratory and require confirmation in other studies. Although age interaction models fit the data better than main effects models, the number of *APOE*-ε4 homozygotes in each age category were relatively small (53 individuals aged 45-54 years; 82 individuals aged 54-65 years; 51 individuals aged 65-74 years; and 29 individuals aged 75-86 years). Overall, the results suggest that *APOE* ε4 allele count is unlikely to be a common cause (confounder) explaining associations between poorer pure-tone audiometric thresholds and poorer cognition or poorer visual acuity and poorer cognition.

The lack of an association between *APOE* ε4 allele count and poorer cognition in the general population argues against a direct mechanism linking the gene to cognitive performance (the “phenotype” hypothesis). The “prodromal” hypothesis (in which the effect of APOE4 on cognition is limited to people who go on to develop Alzheimer’s disease) may still be true.

Cognitive impairment was an exclusion criterion at the time of CLSA cohort recruitment, and individuals in various stages of Alzheimer’s disease are under-represented in the cohort as a consequence. In Canada, the prevalence of dementia more than doubles every 5 years for those age 65 years and older, from less than 1% for those age 65-69 to about 25% for those 85 and older (CIHI, 2018). Thus, dementia (or its prodromal stage of mild cognitive impairment) is likely to be under-represented in older age groups of the CLSA. As time passes and the cohort gets older, our analysis could be repeated but restricted to individuals who were younger at the time of recruitment (e.g., <65 years old), who, from a cognitive perspective, are likely to be more representative of peers in their age group in the general population (because the exclusion criterion would have applied to a very small proportion of people in their age group at the time of recruitment). If the prodromal hypothesis is correct, then*APOE* ε4-related cognitive declines may be observed among those who were younger at the time of recruitment and who go on to develop AD.

It is unclear why ε4 homozygotes aged 54-65 years had *better* executive function than others in their age group. Differences in the association between *APOE* ε4 allele count and cognition have been observed (inconsistently) across the lifespan, but not involving this particular age range.

Some studies have reported significant associations between *APOE* ε4 and better cognition in children and young adults, leading to the *APOE* antagonistic pleiotropy hypothesis (Han and Bondi, 2008; Tuminello and Han, 2011). Pooled effect estimates in two metanalyses, however, were not significant, refuting the hypothesis (Ihle et al., 2012; Weissberger et al., 2018). At the other end of the lifespan, some studies have shown that *APOE* ε4 carriers who are 80 years of age or older demonstrate better cognitive performance than others in their age group (Carrion- Baralt 2009, Duchek et al., 2006). Furthermore, the association between*APOE* ε4 and AD risk is attenuated in the oldest-old (Blacker et al., 1997; Breitner et al., 1999; Corrada et al., 2013).

Based on such findings, researchers have hypothesized that *APOE* ε4 carriers who survive into their 80s and 90s may represent selective survivors resilient to the negative effects of the isotype (Duchek et al., 2006). At least two studies suggest that ε4 may be associated with better cognitive performance among middle aged adults. Gharbi-Meliani *et al* recently reported that among a cohort of 5,561 individuals followed for a mean of 20 years, *APOE*-ε4 heterozygotes had better global cognitive performance than non-ε4 carriers between the ages of 45-55 years, then no differences between ages 60-70 years, and poorer performance from 75 years onwards. The better cognitive performance in the younger *APOE* ε4 heterozygotes was primarily in tests of executive functioning. In contrast, there was no cognitive advantage to *APOE* ε4-homozygosity for any age group, but *APOE* ε4 homozygotes had faster rates of global cognitive decline from age 65 years onwards relative to *APOE*-ε3 homozygotes (Gharbi-Meliani et al., 2021). Jochemson *et al* reported that having one or two *APOE* ε4 alleles was associated with memory increases for persons aged ≤ 57 years, and memory decreases for persons older than 57 years, relative to non-carriers of the same age group over a follow up of 3.8 years (Jochemsen et al., 2012). Our finding of an association between *APOE* ε4 and better executive function among 55-64-year-olds in our study needs to be replicated in other large studies to be considered as more than a sampling error.

This study is the largest to date examining associations between hearing and *APOE* ε4, and the only longitudinal study. It provides evidence that ε4 allele count is not associated with poorer audiometric hearing in older adults and thus cannot be considered a common cause that might underly observed associations between pure-tone thresholds and executive function or memory. Previous studies have been limited by relatively small numbers of participants. The second largest study (Mener et al, 2014) examined 1,833 participants aged 70 years or older in the Health ABC study, and found that a greater number of *APOE* ε4 alleles was associated with marginally better hearing. Only 23 participants (1.3%) were homozygous for*APOE* ε4, raising the possibility of sampling error or volunteer bias (the population prevalence is approximately 2.2%) (Menzel et al, 1983). In the Health ABC study, a higher proportion of Black participants had at least one *APOE* ε4 allele, and the prevalence of hearing loss has been observed to be lower in individuals who are Black (Lin et al, 2012). Kurniawan et al. (2012) examined cross-sectional associations between ε4 allele count and audiometric hearing among 435 participants aged 85 years old from the city of Leiden, Netherlands. Only 6 participants (1.4%) were homozygous for ε4. In contrast to our study and to the Health ABC study, a greater *APOE* ε4 count was associated with worse pure-tone thresholds. It is unclear if the discrepancy in results between studies is due to sampling error, or differences in age ranges or other factors that might modify the strength of the associations.

The lack of association between visual acuity and *APOE* ε4 suggests that the APOE4 isoform is not a common cause of associations between visual acuity and executive function or memory that have been observed in the CLSA (Phillips et al., 2022) and in similar cohorts (Cao et al., 2023). The relationship between *APOE* ε4 and specific eye diseases, while not the focus of our study, appears to be more nuanced; for example, the APOE4 isoform is associated with cataracts, open angle glaucoma and retinal abnormalities, but may protect against the development of age-related macular degeneration (Inoue et al., 2013; Ishida et al., 2004; Utheim et al., 2008).

### 4.1 Threats to internal validity

A substantial number of CLSA participants were missing data for one or more variables used in the analysis. Participants with partially missing data appeared to be different in a number of ways compared to participants with complete data, based on available information. On average, they were older and unhealthier according to a number of metrics, but the groups did not differ in terms of distribution of *APOE* ε4 alleles, suggesting that the health differences between groups with missing and complete data were not driven byε4 isoform. Still, 9,558 of the 16,469 participants with missing data (58%) were missing data on *APOE* genotype, and so the true distribution of *APOE* ε4 isotype among the group with missing data is unknown.

We considered the possibility that individuals with *APOE* ε4 alleles would be less likely to join the CLSA because they were more likely to have Alzheimer’s disease or other*APOE* ε4-related health problems (including, potentially, cognitive impairment, hearing loss or vision loss) that would make it difficult to participate. If so, then the proportion of CLSA participants with*APOE* ε4 alleles might be expected to decrease as a function of age since most *APOE* ε4-related health conditions increase in prevalence with age. This was not the case. Furthermore, allelic frequencies in the analytic sample (ε2: 8.2%; ε3: 77.8%; ε4: 14.1%) are very similar to previously published estimates, suggesting that individuals with the *APOE* ε4 allele were not under- represented in the CLSA sample. For example, Farrer et al, in a 1997 meta-analysis, estimated that the worldwide frequencies of ε2, ε3 and ε4 were 8.4%, 77.9% and 13.7%, respectively (Farrer et al, 1997). To reduce the risk of both attrition and volunteer bias, we adjusted forε4- related health conditions in multivariable models, but doing so did not significantly alter effect estimates versus unadjusted models. Thus, it appears unlikely that volunteer bias (in which only “healthy” individuals with the *APOE* ε4 allele participated in the study) can explain the negative results.

We chose to analyze audiometric hearing, pinhole-corrected visual acuity and five specific measures of cognition (combined into two principal component scores) because they were measured in the CLSA and are commonly used in studies assessing associations between sensory loss and cognition. Pinhole-corrected visual acuity was analyzed rather than habitually corrected visual acuity because there is no evidence to suggest that refractive error is a function of *APOE*. Future studies might evaluate associations between *APOE* and measures of supra-threshold auditory processing (e.g., speech-in-noise understanding), or between*APOE* and visual contrast sensitivity, which is more sensitive to age-related changes than visual acuity (Eisner et al., 1987; Schneck et al., 2004). Other genetic or epigenetic mechanisms remain to be investigated.

## 5. Conclusions

In a large population-based study, *APOE* ε4 allele count was not associated with baseline differences or 3-year changes in executive function, memory, pure-tone hearing thresholds, or visual acuity. Thus, the *APOE* ε4 gene variant is unlikely to be a common cause explaining associations between audiometric hearing loss and declines in executive function or memory, or between poor visual acuity and declines in executive function or memory. Other, unknown age-related factors may underlie the associations,(Christensen et al., 2001; Lindenberger and Baltes, 1997) or it may be that there is a (yet unproven) causal link between sensory loss and cognitive decline.

## Data Availability

Data are available from the Canadian Longitudinal Study on Aging (www.clsa-elcv.ca) for researchers who meet the criteria for access to de-identified CLSA data.

## Acknowledgements

This particular analysis was funded by a CLSA Catalyst Grant awarded by the Canadian Longitudinal Study on Aging (Funding Reference Number (FRN): 170301).

Drs. Mick, Phillips, Pichora-Fuller, and Wittich are members of Team 17 (Interventions at the Sensory-Cognitive Interface), and Dr. Gan-or is a member of Team 1 (Clinical Genetics and Gene Discovery), of the Canadian Consortium on Neurodegeneration in Aging (CCNA). The Canadian Consortium on Neurodegeneration in Aging is supported by a grant from the Canadian Institutes of Health Research with funding from several partners.

This research was made possible using the data/biospecimens collected by the Canadian Longitudinal Study on Aging (CLSA). Funding for the Canadian Longitudinal Study on Aging (CLSA) is provided by the Government of Canada through the Canadian Institutes of Health Research (CIHR) under grant reference: LSA 94473 and the Canada Foundation for Innovation. This research has been conducted using the CLSA Baseline Comprehensive Dataset version 4.1, Follow-up 1 Comprehensive Dataset version 3.0, and the Vital Status files for both Tracking and Comprehensive cohorts, under Application Number 20CA007. The CLSA is led by Drs. Parminder Raina, Christina Wolfson and Susan Kirkland.

Data are available from the Canadian Longitudinal Study on Aging (www.clsa-elcv.ca) for researchers who meet the criteria for access to de-identified CLSA data. The opinions expressed in this manuscript are the author’s own and do not reflect the views of the Canadian Longitudinal Study on Aging.

## Author contributions

Mick: Concepualization, data curation, formal analysis; funding acquisition; investigation; methodology; project administration; resources; supervision; validation; visualization; Writing: original draft, review and editing.

Kabir: Data curation, formal analysis; project administration; Writing: original draft, review and editing.

Karunatilake: Writing: original draft, review and editing.

Phillips: Concepualization; funding acquisition; investigation; methodology; project administration; resources; supervision; Writing: review and editing.

Pichora-Fuller: Concepualization; funding acquisition; investigation; methodology; project administration; resources; supervision; Writing: review and editing.

Sosero: Formal analysis; Writing: review and editing.

Gan-or: Formal analysis; supervision; Writing: review and editing.

Wittich: Concepualization; funding acquisition; investigation; methodology; project administration; resources; supervision; Writing: review and editing.

